# Seroprevalence of SARS-CoV-2 among adults in three regions of France following the lockdown and associated risk factors: a multicohort study

**DOI:** 10.1101/2020.09.16.20195693

**Authors:** Fabrice Carrat, Xavier de Lamballerie, Delphine Rahib, Hélène Blanché, Nathanael Lapidus, Fanny Artaud, Sofiane Kab, Adeline Renuy, Fabien Szabo de Edelenyi, Laurence Meyer, Nathalie Lydié, Marie-Aline Charles, Pierre-Yves Ancel, Florence Jusot, Alexandra Rouquette, Stéphane Priet, Paola Mariela Saba Villarroel, Toscane Fourié, Clovis Lusivika-Nzinga, Jerome Nicol, Stephane Legot, Nathalie Druesne-Pecollo, Younes Esseddik, Cindy Lai, Jean-Marie Gagliolo, Jean-François Deleuze, Nathalie Bajos, Gianluca Severi, Mathilde Touvier, Marie Zins, for the SAPRIS and SAPRIS-SERO study groups

**Affiliations:** Sorbonne Université, Inserm, Institut Pierre-Louis d’Epidémiologie et de Santé Publique, Paris, France; Département de Santé Publique, APHP.Sorbonne Université, Paris, France; Unité des Virus Emergents, UVE: Aix Marseille Univ, IRD 190, INSERM 1207, IHU Méditerranée Infection, 13005, Marseille, France; Santé publique France, Saint-Maurice, France; Fondation Jean Dausset-CEPH (Centre d’Etude du Polymorphisme Humain), CEPH-Biobank, Paris, France; CESP UMR1018, Université Paris-Saclay, UVSQ, Inserm, Gustave Roussy, Villejuif, France; Paris University, Paris, France; Paris Saclay University, Inserm UMS 11 Villejuif, France; Sorbonne Paris Nord University, Inserm U1153, Inrae U1125, Cnam, Nutritional Epidemiology Research Team (EREN), Epidemiology and Statistics Research Center – University of Paris (CRESS), Bobigny, France; Université Paris Saclay, Inserm, CESP U1018, Le Kremlin Bicêtre, France; Service de Santé Publique, APHP.Paris Saclay, Le Kremlin Bicêtre, France; Ined, Inserm, EFS, UMS Elfe, Aubervilliers France; Obstetrical, Perinatal and Pediatric Epidemiology Research Team, Center for Epidemiology and Statistics Sorbonne Paris Cité, INSERM U1153, Paris Descartes University, Paris, France; Clinical Research Unit, Center for Clinical Investigation P1419, Cochin Broca Hôtel-Dieu Hospital, Paris, France; Université Paris-Dauphine, PSL-Research University, LEDa, Paris, France; Institut de santé publique, Pôle recherche clinique, Institut national de la santé et de la recherche médicale, Paris, France; Inserm, institut thématique santé publique, Paris, France; IRIS, Inserm/EHESS/CNRS, Aubervilliers, France; Department of Statistics, Computer Science and Applications, University of Florence, Italy

**Keywords:** SARS-COV-2, COVID-19, general population, cohort, seroprevalence, risk factors

## Abstract

**Background:** To estimate the seroprevalence of SARS-CoV-2 infection in May-June 2020 after the lockdown in adults living in three regions in France and to identify the associated risk factors.

**Methods:** Participants in a survey on COVID-19 from an existing consortium of three general adult population cohorts living in the Ile-de-France (IDF) or Grand Est (GE) - two regions with high rate of COVID-19, or in the Nouvelle-Aquitaine (NA) – with a low rate, were asked to take a dried-blood spot (DBS) for anti-SARS-CoV-2 antibodies assessment.

The primary outcome was a positive anti-SARS-CoV-2 ELISA IgG result against the spike protein of the virus (ELISA-S). The secondary outcomes were a positive ELISA IgG against the nucleocapsid protein (ELISA-NP), anti-SARS-CoV-2 neutralizing antibodies titers ≥40 (SN), and predicted positivity obtained from a multiple imputation model (MI). Prevalence estimates were adjusted using sampling weights and post-stratification methods.

**Findings:** Between May 4, 2020 and June 23, 2020, 16,000 participants were asked to provide DBS, and 14,628 were included in the analysis, 983 with a positive ELISA-S, 511 with a positive ELISA-NP, 424 with SN≥40 and 941±31 with a positive MI. Adjusted estimates of seroprevalence (positive ELISA-S) were 10.0% (95%CI 9.1%;10.9%) in IDF, 9.0% (95%CI 7.7%; 10.2%) in GE and 3.1% (95%CI 2.4%; 3.7%), in NA. The adjusted prevalence of positive ELISA-NP, SN and MI were 5.7%, 5.0% and 10.0% in IDF, 6.0%, 4.3% and 8.6% in GE, and 0.6%, 1.3% and 2.5% in NA, respectively. A higher seroprevalence was observed in younger participants and when at least one child or adolescent lived in the same household. A lower seroprevalence was observed in smokers compared to non-smokers.

**Interpretation:** At the end of the lockdown the prevalence of anti-SARS-CoV-2 IgG or neutralizing antibodies remained low in the French adult population, even in regions with high reported rates of COVID-19.

## Introduction

Serological surveys help determine the extent of infection by a viral agent in a population and identify associated risk factors.^1^ In addition, characterizing the distribution of antibodies against this agent can help evaluate the portion of the population that is immunized to quantify herd immunity. However, despite the ongoing COVID-19 pandemic, there are still very few serologic surveys describing the factors associated with SARS-CoV-2 seroprevalence, and only one study explored the distribution of neutralizing antibodies against SARS-CoV-2 in a general adult population in a very low prevalence area.^2^

Serologic surveys of SARS-CoV-2 have been performed between January 2020 and July 2020 in the general population in Iceland,^3^ Switzerland,^4^ Spain,^5^ UK,^6,7^ Italy,^8^ Belgium,^9^ Germany,^2^ China,^10^ Brazil,^11^ Canada,^12^ and the US.^13-15^ They all showed a low seroprevalence in the general population (<10%), and sometimes identified associations between a positive test result and younger age, sex, ethnicity, as well as lower socioeconomic status and population density.

In France, SARS-CoV-2 positive RT-PCR tests were first reported in imported cases on week 4 (January 24, 2020), generalized lockdown began on week 12 (March 17, 2020) and emergency room visits for possible COVID-19 peaked on week 13, decreasing thereafter. This led the French government to ease lockdown restrictions on week 20 (May 11, 2020).

Our main goals were 1) to estimate the seroprevalence of SARS-CoV-2 infection in the French adult population at the end of lockdown in three regions and 2) to identify the associated risk factors.

## Participants and Methods

### Design

The present report combined data collected from questionnaires in the SAPRIS (“SAnté, Perception, pratiques, Relations et Inégalités Sociales en population générale pendant la crise COVID-19”) survey in France, with serological results from the SAPRIS-SERO study.

The SAPRIS survey has been described elsewhere.^16^ Briefly, the survey was created in March 2020 to evaluate the main epidemiological, social and behavioral challenges of the SARS-CoV2 epidemic in France in relation to social inequalities in health and healthcare. It is based on a consortium of prospective cohort studies including three general population-based adult cohorts and two child-cohorts (not presented in this study).^17-19^ All participants from the original cohorts with regular access to electronic (internet) questionnaires were invited to participate in the SAPRIS survey (supplementary figure 1). Two self-administered questionnaires covering the lockdown and the post-lockdown periods were sent as of April 1, 2020 and returned before May 27, 2020. Variables collected in the questionnaires included socio-demographics, household size and composition, COVID-19 diagnosis, SARS-CoV-2 RT-PCR test, a detailed description of the subject’s symptoms in the two weeks before each questionnaire, comorbidities, healthcare use and treatment, employment, daily life, child care, alcohol, tobacco and cannabis use, social and sexual life, preventive measures, risk perception and beliefs. The goal of the SAPRIS-SERO study (#NCT04392388) including participants enrolled in the SAPRIS survey, was to quantify and follow the cumulative incidence of SARS-CoV-2 infection in the French population using serological tests and to assess the determinants of infection. Self-sampling dried-blood spot (DBS) kits were mailed to each participant including material (a DBS card, lancets, pad), detailed printed instructions on how to perform the test, and a self-addressed stamped padded envelope to be returned with the card to the centralized biobank (CEPH Biobank, Paris, France). Kits were received, then blood spots were visually assessed and registered in the CEPH-Biobank LIMS (BIOBASE). Four 4.7 mm discs were punched of the spots on Panthera™ (PerkinElmer) and stored in 2D FluidX 96-Format 0.5 mL tubes (Brooks) in -30°C freezers. Tubes were sent to the virology laboratory (Unité des virus Émergents, Marseille, France) for serological analysis. Eluates were processed with a commercial Elisa test (Euroimmun®, Lübeck, Germany) to detect anti-SARS-CoV-2 antibodies (IgG) directed against the S1 domain of the spike protein of the virus (ELISA-S). The volume of eluate used corresponded to the amount of serum and dilution recommended in the manufacturer’s instructions. All samples with an ELISA-S test optical density ratio ≥ 0.7 were also tested with an ELISA test to detect IgG antibodies against the SARS-CoV-2 nucleocapsid protein (Euroimmun®, Lübeck, Germany, ELISA-NP) and with an in-house micro-neutralization assay to detect neutralizing anti-SARS-CoV-2 antibodies (SN), as described elsewhere.^20^ Briefly, we used VeroE6 cells cultured in 96-well microplates, 100 TCID50 of the SARS-CoV-2 strain BavPat1 (courtesy of Pr. Drosten, Berlin, Germany) and serial dilutions of serum (1/20–1/160). Dilutions associated with the presence or absence of a cytopathic effect on post-infection day 4.5 were considered to be negative or positive, respectively. The neutralization titer referred to the highest dilution of positive serum. The specificity of the assay was close to 100% in a population of blood donors sampled in 2017-2018 when samples with a titer ≥40 were considered to be positive.^20^

We randomly selected 16,000 of the participants of the SAPRIS survey for this study who agreed to be tested and who were residents from one of the three French administrative regions: Ile-de-France (IDF) or Grand Est (GE), i.e. the two regions with the highest reported cumulated rates of COVID-19 at the end of the lockdown period, or Nouvelle-Aquitaine (NA), a region with a low reported rate.^21^

Ethical approval and written or electronic informed consent were obtained from each participant before enrolment in the original cohort. The SAPRIS survey was approved by the Inserm ethics committee (approval #20-672 dated March 30, 2020). The SAPRIS-SERO study was approved by the Sud-Mediterranée III ethics committee (approval #20.04.22.74247) and electronic informed consent was obtained from all participants for DBS testing.

### Outcomes

The main outcome was a positive ELISA-S test. In accordance with the manufacturer’s instructions a test was considered to be ELISA-positive with an optical density ratio ≥ 1.1, ELISA-indeterminate between 0.8 and 1.1, and ELISA-negative, <0.8. The secondary outcomes were a positive ELISA-NP (using the same thresholds) and positive SN defined as a titer ≥40. Because test sensitivity and specificity was not 100%, we also used a multiple imputation (MI) method to estimate a participant’s positivity in which the likelihood of positivity was based on observed test results and covariates.

### Covariates

The association of seroprevalence was evaluated in relation to age, gender, socio-demographic characteristics, BMI, chronic conditions (according to a pre-specified list), tobacco and alcohol use before the lockdown. Age groups were categorized according to predefined limits (<40; 40-49; 50-59; 60-69; ≥70 years old) and BMI according to standard cut-offs (<18.5; 18.5-<25; ≥25-<30; ≥30 kg/m^2^).^22^ The association of seroprevalence was also studied in relation to symptoms. Possible COVID-19 was defined according to the European Centre for Disease Prevention and Control as at least one of the following: cough, fever, dyspnea, and sudden anosmia, ageusia or dysgeusia.^23^ Participants who did not report any of these symptoms on either questionnaire, did not have a positive COVID-19 diagnosis, or did not experience cough, fever or feverishness from the beginning of the year were considered to be asymptomatic.

### Statistical methods

Inverse probability weighting and generalized raking were used to estimate seroprevalence in the adult population.^24^ Weights were estimated from each cohort source by logistic regression, with selection or participation as response variables and socio-demographic characteristics as covariates. An initial cohort-specific calibration was performed by generalized raking in relation to the marginal totals of the distribution of age class, gender and socio-professional category in the target population. The weights were then rescaled according to the relative sample size of each cohort, then recalibrated according to the same covariates to provide representative estimates of the adult population. This weighting procedure was performed for each region independently. Confidence intervals for weighted estimates were computed by bootstrapping.

To fit the MI model, participants with all three positive ELISA-S, ELISA-NP and SN test results were classified as “true” positives while those with all three negative results or ELISA-S <0.7 were “true” negatives. The Markov Chain Monte Carlo method was used to imputing MI using numerical values from the three serological tests (log-transformed), region, age and gender. The MI was built from 100 imputed data sets and estimates combined with Rubin’s rules.^25^

Chi-Square test for trend was used on unweighted data to compare symptoms and health care use according to ELISA-S results. Logistic regression models were used on unweighted data with stratification in the source cohort to identify the determinants of a positive ELISA-S (primary outcome). Indeterminate ELISA-S results were grouped with negative results in the primary analysis. Multivariable analysis was performed including region, age, gender and all factors associated with seroprevalence in univariable analysis. A backward elimination procedure was used to identify independent covariates associated with a positive ELISA-S. Contact with a RT-PCR positive household member was not considered to prevent the risk of reverse causation. Multivariable analyses were repeated using secondary outcomes then performed in each region to identify any potential regional effect-modification.

Weighting and multiple imputation used the survey and mice package from R software version 3.6.3 (R Foundation for Statistical Computing, Vienna, Austria). Other analyses were performed with SAS 9·4 software (SAS Institute Inc., Cary, North Carolina, USA). P<.05 was considered to be statistically significant.

## Results

A total of 116,903 out of 279,478 (42%) adults who were invited to participate in the survey completed the 1^st^ questionnaire and 108,595 (39%) the 2^nd^ questionnaire (94,999 completed both) while 104,001 (80% of participants to the 1^st^ or 2^nd^ questionnaire) accepted the serological study (supplementary figure 1). Sixteen thousands of the 36,531 participants living in IDF, GE or NA who agreed to the serological study and had the 1^st^ and 2^nd^ questionnaire validated were invited to perform the DBS. The DBS was returned by 15,414 (96%) of these participants and serology was performed on 14,830 (93%) samples: 14,628 (91%) could be interpreted and were included in the analyses. The median time between the 2^nd^ questionnaire and DBS was 12 days (Q1-Q3: 10-16 days). Ninety percent of DBS samples were performed at the end of lockdown between weeks 19 and 21 (May, 4 - May, 24 2020). Participant characteristics are described in supplementary table 1.

### Seroprevalence of SARS-CoV-2

Nine hundred eighty-three participants had a positive ELISA-S, 552 in IDF, 270 in GE and 161 in NA, with weighted seroprevalence estimates in the adult population of 10.0% in IDF (95%CI 9.1%; 10.9%), 9.0% in GE (95% CI 7.7%; 10.2%) and 3.1% in NA (95% CI 2.4%; 3.7%) (table 1). The seroprevalence estimates of positive ELISA-NP and SN were markedly lower: 5.7% and 5.0%, in IDF, 6.0% and 4.3% in GE, and 0.6% and 1.3% in NA, respectively. Two hundred ninety-two participants were positive with all three methods and 13,314 were negative with all methods or had an ELISA-S <0.7, while the MI was used to predict 1,022 (supplementary figure 2): 941 participants (SD=31) were classified as MI positive, 548 (SD=23) in IDF, 259 (SD=16) in GE, 134 (SD=12) in NA, with weighted MI seroprevalence estimates in the adult population of 10.0% in IDF (95%CI 8.9%; 11.3%), 8.6% in GE (95%CI 7.2%; 10.4%) and 2.5% in NA (95%CI 1.9%; 3.3%). The sensitivity and specificity of ELISA-S in relation to MI was 97.9% (95% CI 96.9%: 98.9%) and 97.7% (95%CI 97.4%; 98.0%), respectively. The sensitivity and specificity of ELISA-NP was 50.3% (95%CI 46.9%; 53.6%) and 99.5% (95%CI 99.3%; 99.6%), respectively, and that of SN was 41.4% (95% CI 38.2%; 44.7%) and 99.5% (95%CI 99.3%; 99.6%), respectively.

**Table 1.** Weighted prevalence estimates.

|  |  | ELISA-S | ELISA-NP | SN | MI |
| --- | --- | --- | --- | --- | --- |
| Ile-de-France<br>(IDF) - N=6,348 | number positive<br>weighted prevalence<br>(95% CI) | 552<br>10.0%<br>(9.1% ; 10.9%) | 315<br>5.7%<br>(4.9% ; 6.4%) | 253<br>5.0%<br>(4.2% ; 5.7%) | 548±23<br>10.0%<br>(8.9% ; 11.3%) |
| Grand Est<br>(GE) - N=3,434 | number positive<br>weighted prevalence<br>(95% CI) | 270<br>9.0%<br>(7.7% ; 10.2%) | 161<br>6.0%<br>(4.9% ; 7.0%) | 120<br>4.3%<br>(3.2% ; 5.2%) | 259±16<br>8.6%<br>(7.2% ; 10.4%) |
| Nouvelle-Aquitaine (NA)<br>N=4,846 | number positive<br>weighted prevalence<br>(95% CI) | 161<br>3.1%<br>(2.4% ; 3.7%) | 35<br>0.6%<br>(0.3% ; 0.9%) | 51<br>1.3%<br>(0.8% ; 1.7%) | 134±12<br>2.5%<br>(1.9% ; 3.3%) |

### Symptoms and healthcare use

Participants with a positive ELISA-S had a higher rate of self-reported symptoms than those with negative tests except for skin lesions (table 2). Forty-seven percent of ELISA-S positive participants experienced symptoms of possible COVID-19 a median of 56 days (Q1: 40 days; Q3: 61 days) before collection of blood samples, while the rate was 24% at 53 days (46, 60) in those with indeterminate results, and 19% at 53 days (42, 61) in participants with negative results (P<0.0001). ELISA-S was positive in 74% (95%CI 64%; 84%) of participants with a positive RT-PCR test, in 47% (95%CI 43%; 51%) of those with anosmia or ageusia, in 44% (95%CI 40%; 48%) with a medical diagnosis of COVID-19, in 15% (95% CI 14%; 17%) with symptoms of possible COVID-19, and in 3.7% (95% CI 3.2%; 4.2%) of asymptomatic participants. The proportion of positive ELISA-S in participants with possible COVID-19 was higher in IDF and GE than in NA (figure 1). It also varied during lockdown and decreased from 25% in IDF (95%CI 21%; 29%), 26% in GE (95%CI 20%; 32%) and 5.3% in NA (95%CI 2.5%; 9.9%) when the onset of possible COVID-19 symptoms were reported during week 12 (March 16 - 22 – the beginning of lockdown) to 2.7% in IDF (95%CI 0.1%; 14%), 0.0% in GE (95%CI 0%; 23%) and 2.9% in NA (0.1% 15.3%), when the onset of symptoms was reported during week 18 (figure 1).

**Figure 1.**
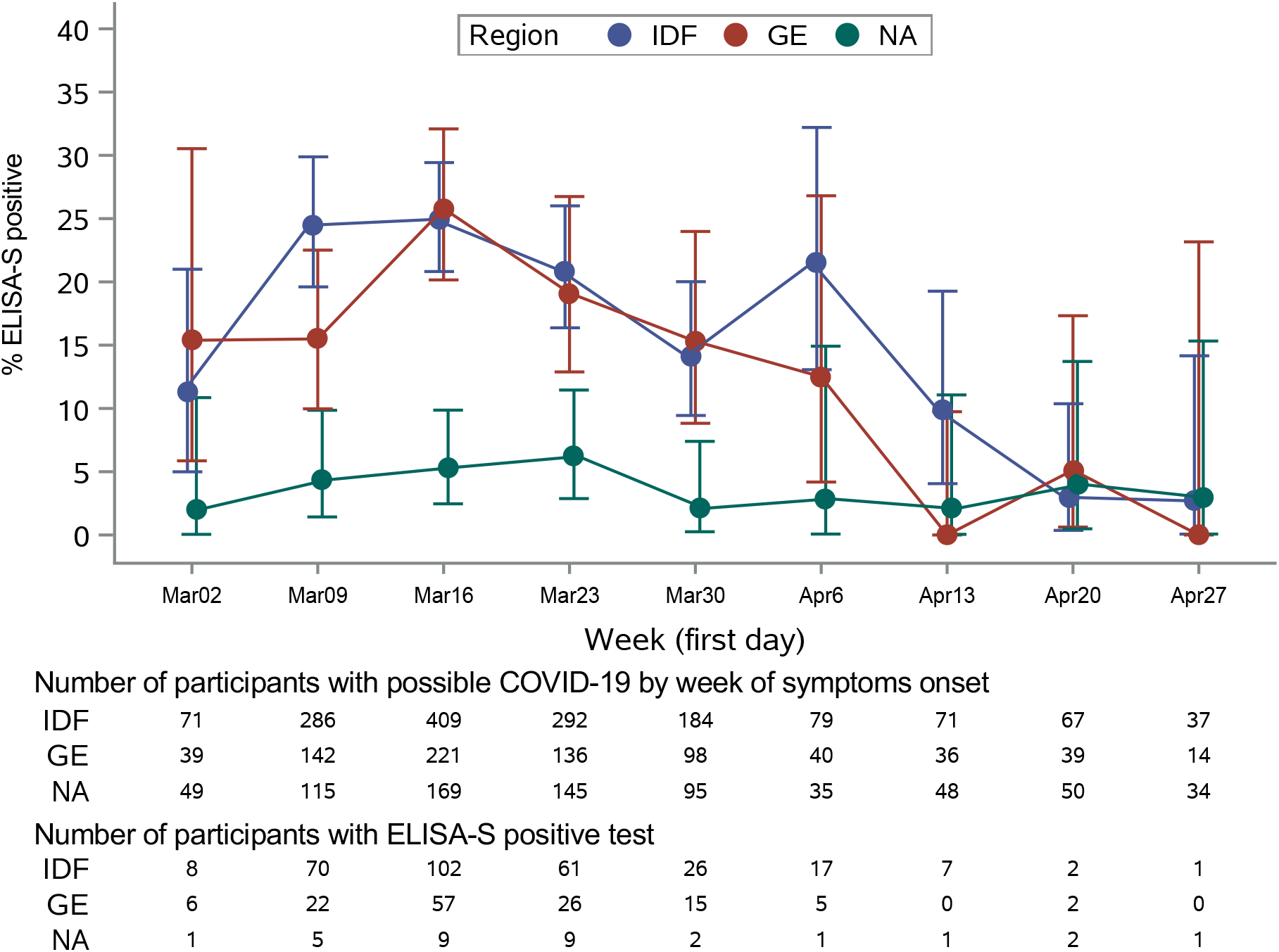
Proportion of participants with possible COVID-19 and a positive ELISA-S serological result according to the date of the onset of symptoms.

**Table 2.**
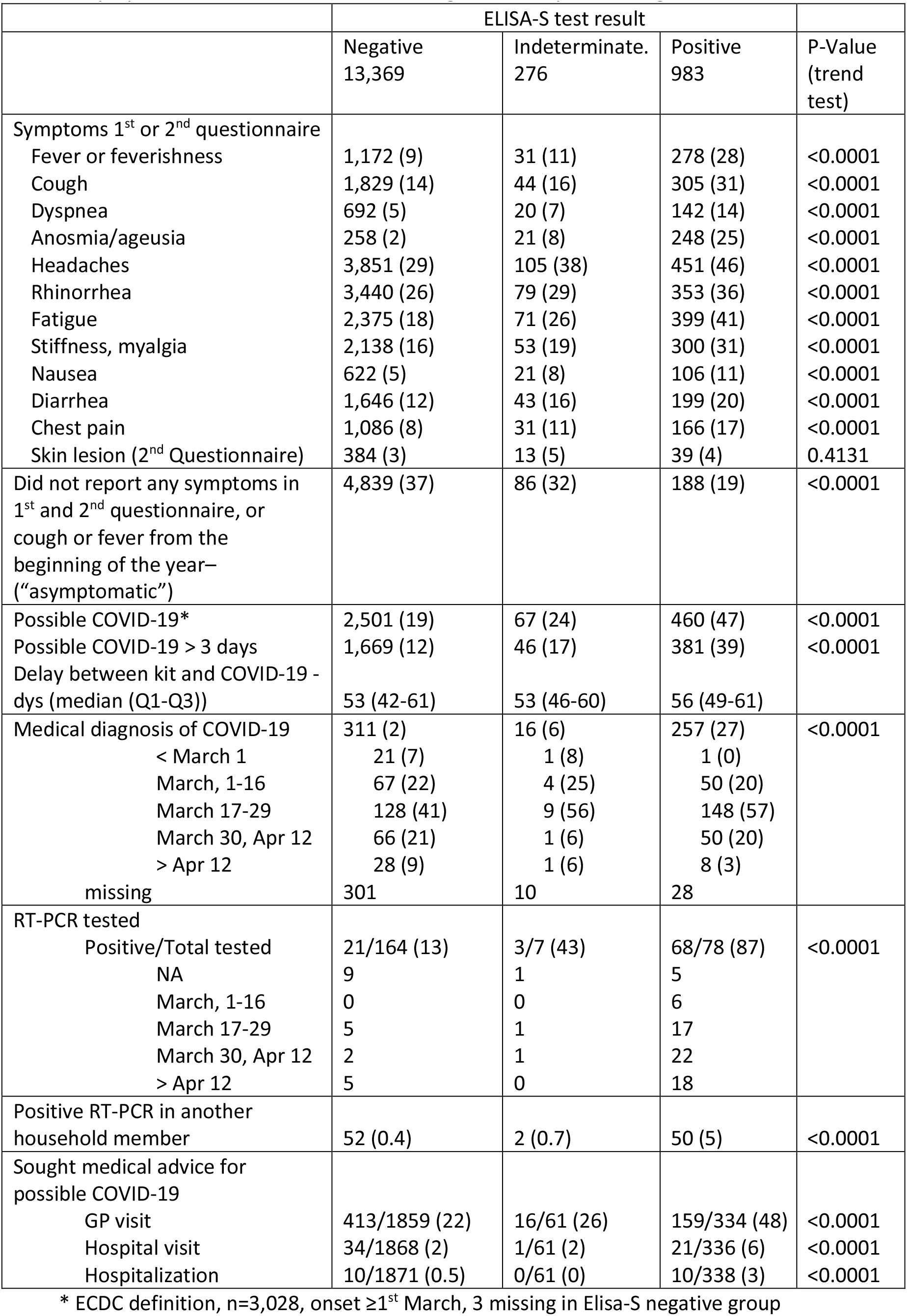
Symptoms and health care use during the survey according to ELISA-S results

In participants with a positive ELISA-S, a positive ELISA-NP was found in 29/185 (16%) asymptomatic participants, in 335/454 (74%) with possible COVID-19, and in 88/319 (28%) who reported other symptoms (P<0.0001, supplementary figure 3), while a positive SN was found in 40/188 (21%), 250/459 (54%) and 81/322 (25%), respectively (P<0.0001).

### Factors associated with seroprevalence

On univariable analysis, the rate of positive ELISA-S was higher in IDF and GE than in NA (table 3) as well as in younger adult groups with an observed peak in ages 35 to 44 years old in each region (figure 2). The association with age was similar with positive MI, although a higher proportion of positive SN or ELISA-NP was observed in the youngest age groups in the IDF and NA regions (supplementary figures 4&5). Multivariable analysis showed an independent and positive association between positive ELISA-S, IDF and GE compared to NA, for younger age, and at least one child or adolescent living in the same household (table 4). A negative association was found with active smoking (*vs* no smoking). The observed associations were confirmed with MI and were overall consistent with ELISA-NP and SN, although they did not all reach statistical significance due to a smaller number of events (supplementary tables 2-4). When multivariable analysis was performed in each region separately, the associations did not differ between IDF and GE but the pattern in NA was different from that in IDF or GE, with a higher Odds-Ratio (OR) in young age groups in the former (OR= 3.30 (95%CI 1.79; 6.09) in <40 *vs* [50-60] and OR= 3.89 (95%CI 2.18; 6.95) in [40-50] *vs* [50-60]) than in IDF (OR=1.80 (95%CI 1.37; 2.38) and OR=1.83 (95%CI 1.39; 2.40), respectively) and GE (OR=1.45 (95%CI 0.97; 2.17) and OR=1.44 (95%CI 0.99; 2.11), respectively). Moreover, there was a significant association with female gender in NA (OR=2.11 (95%CI 1.42; 3.14)) but not in IDF (OR= 1.00 (95%CI 0.83; 1.22)) or GE (OR=1.02 (95%CI 0.76; 1.36)) (figure 3).

**Figure 2.**
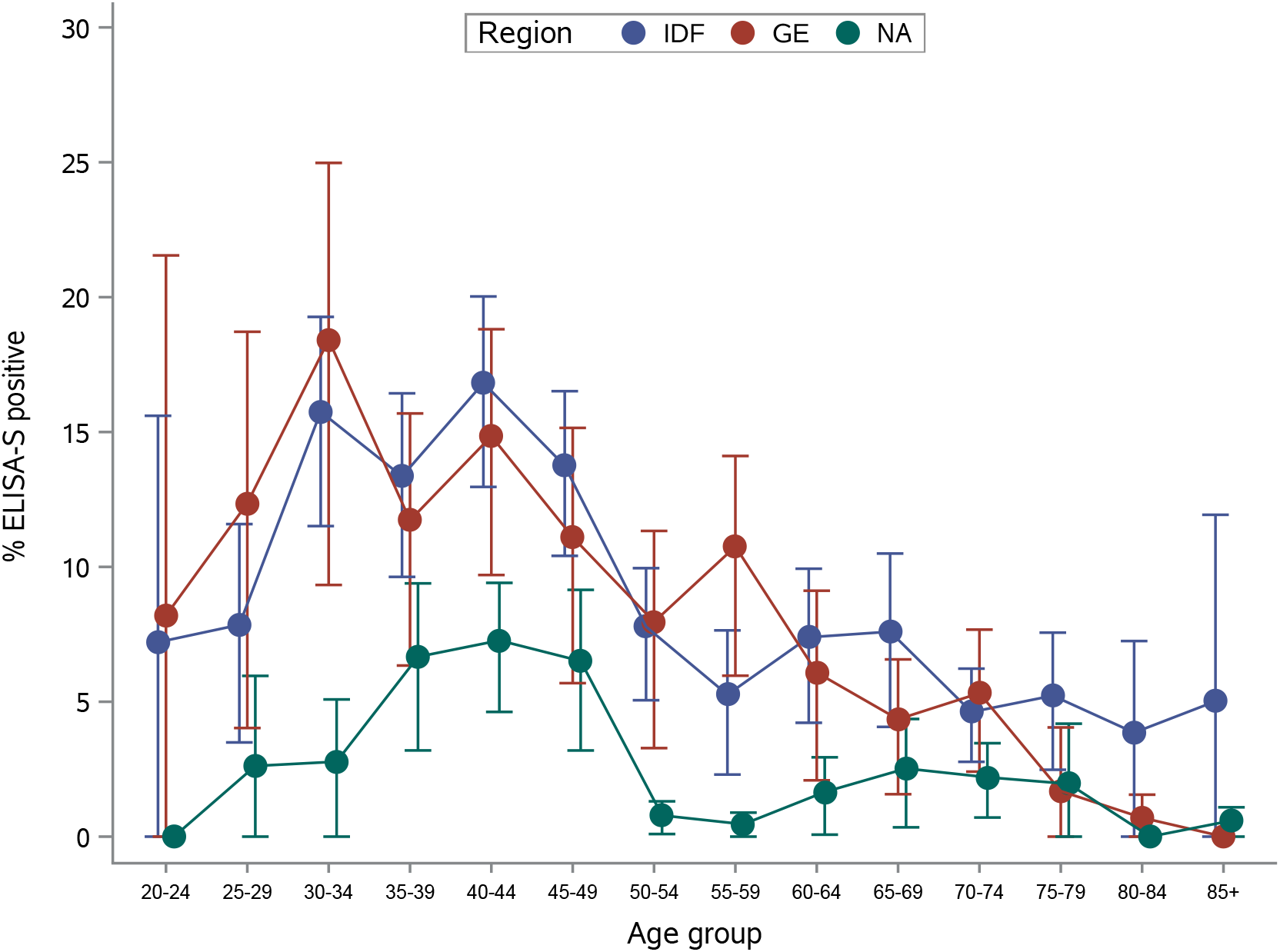
Proportion of participants with a positive ELISA-S by age (weighted estimates).

**Figure 3.**
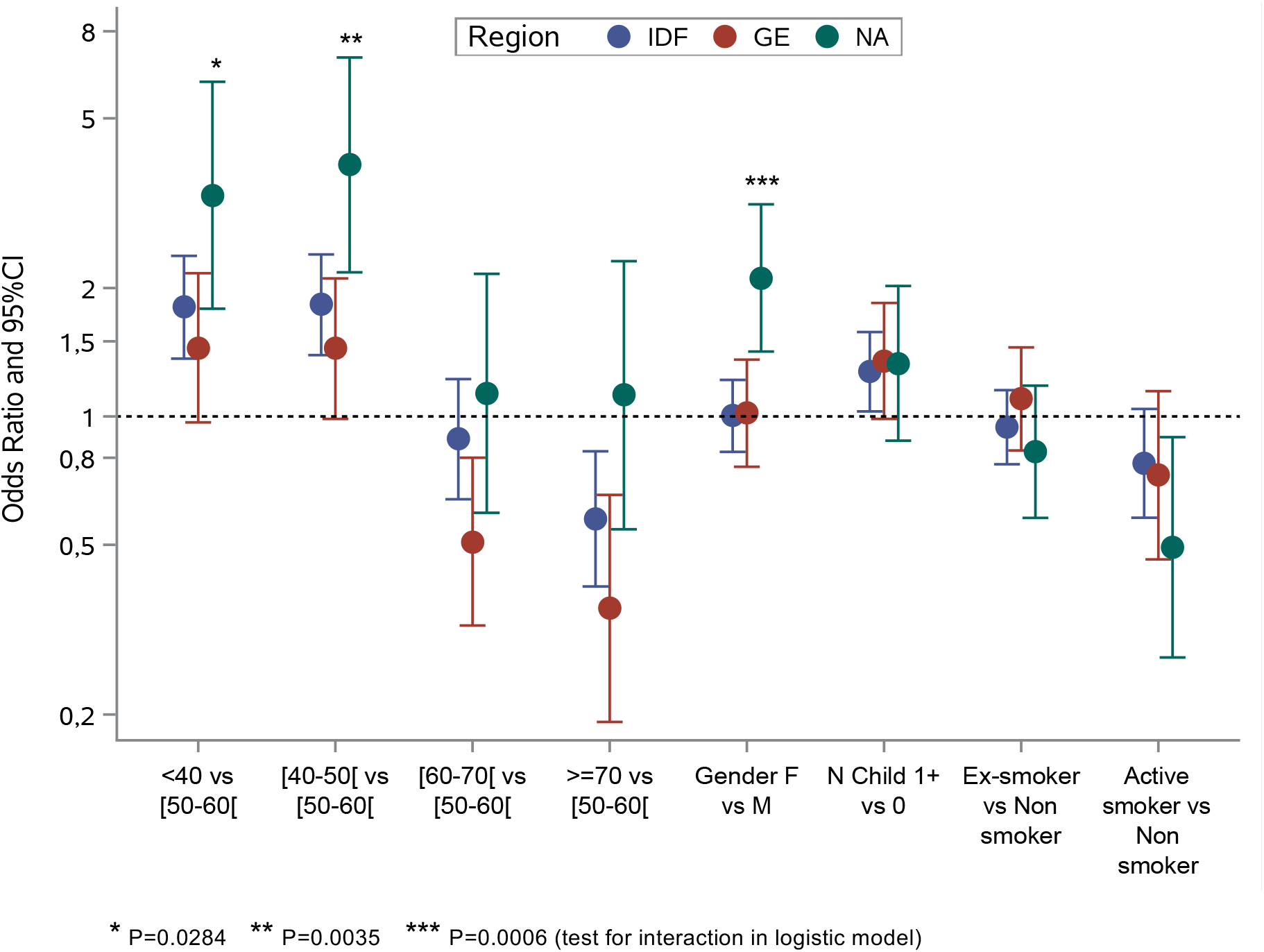
Risk factors of positive ELISA-S by French region.

**Table 3.** Factors associated with a positive ELISA-S (vs negative or indeterminate)

|  | Positive/Total | % (Exact 95%CI) | Weighted prevalence estimates (%) | Odds-Ratio* | 95%CI | P-value |
| --- | --- | --- | --- | --- | --- | --- |
| Regions |  |  |  |  |  | <0.0001 |
| Ile-de-France | 552/6,348 | 8.7 (8.0 ; 9.4) | 10.0 (9.1 ; 10.9) | 2.64 | 2.20 ; 3.17 | <0.0001 |
| Grand-Est | 270/3,434 | 7.9 (7.0 ; 8.8) | 9.0 (7.7 ; 10.2) | 2.44 | 1.99 ; 2.98 | <0.0001 |
| Nouvelle Aquitaine | 161/4,846 | 3.3 (2.8 ; 3.9) | 3.1 (2.4 ; 3.7) | Reference |  |  |
| Age group (years) |  |  |  |  |  | <0.0001 |
| <40 | 245/2,262 | 10.8 (9.6 ; 12.2) | 10.9 (9.2 ; 12.4) | 2.06 | 1.68 ; 2.53 | <0.0001 |
| [40-50] | 332/2,897 | 11.5 (10.3 ; 12.7) | 12.9 (11.2 ; 14.5) | 2.14 | 1.77 ; 2.59 | <0.0001 |
| [50-60] | 176/3,019 | 5.8 (5.0 ; 6.7) | 5.6 (4.3 ; 6.8) | Reference |  |  |
| [60-70] | 133/3,272 | 4.1 (3.4 ; 4.8) | 5.2 (4.0 ; 6.4) | 0.68 | 0.54 ; 0.86 | <0.0001 |
| >=70 | 97/3,175 | 3.1 (2.5 ; 3.7) | 3.8 (2.9 ; 4.6) | 0.50 | 0.38 ; 0.65 | <0.0001 |
| Gender |  |  |  |  |  | 0.0012 |
| Male | 327/5,809 | 5.6 (5.1 ; 6.3) | 7.2 (6.2 ; 8.2) | Reference |  |  |
| Female | 656/8,818 | 7.4 (6.9 ; 8.0) | 8.6 (7.7 ; 9.4) | 1.27 | 1.10 ; 1.46 |  |
| Living Area |  |  |  |  |  | 0.0006 |
| Rural | 118/2,176 | 5.4 (4.5 ; 6.5) | 7.1 (5.6 ; 8.5) | Reference |  |  |
| <20,000 inhab. | 129/1,863 | 6.9 (5.8 ; 8.2) | 8.9 (7.1 ; 10.5) | 1.23 | 0.95 ; 1.60 | 0.1124 |
| 20-000-100,000 inhab. | 211/2,797 | 7.5 (6.6 ; 8.6) | 8.0 (6.6 ; 9.3) | 1.34 | 1.06 ; 1.70 | 0.0135 |
| >100,000 inhab. | 524/7,769 | 6.7 (6.2 ; 7.3) | 8.0 (7.2 ; 8.8) | 1.59 | 1.27 ; 1.99 | <0.0001 |
| Missing | 23 |  |  |  |  |  |
| Household size and composition |  |  |  |  |  | <0.0001 |
| Nb adults (inc. participant) |  |  |  |  |  |  |
| 1 | 176/2,851 | 6.2 (5.3 ; 7.1) | 7.2 (5.8 ; 8.4) | Reference |  |  |
| 2 | 329/6,533 | 5.0 (4.5 ; 5.6) | 6.6 (5.7 ; 7.3) | 0.84 | 0.69 ; 1.01 | 0.0685 |
| 3+ | 478/5,244 | 9.1 (8.4 ; 9.9) | 10.3 (9.1 ; 11.4) | 1.55 | 1.29 ; 1.86 | <0.0001 |
| Nb children (<18yrs) |  |  |  |  |  | <0.0001 |
| 0 | 574/10,848 | 5.3 (4.9 ; 5.7) | 6.7 (6.0 ; 7.4) | Reference |  |  |
| 1+ | 409/3,780 | 10.8 (9.9 ; 11.9) | 11.8 (10.3 ; 13.2) | 2.16 | 1.89 ; 2.48 |  |
| Nb rooms |  |  |  |  |  | 0.0069 |
| 1-2 | 131/1,696 | 7.7 (6.5 ; 9.1) | 9.5 (7.8 ; 11.0) | 1.06 | 0.86 ; 1.29 | 0.6076 |
| 3-4 | 421/5,715 | 7.4 (6.7 ; 8.1) | 8.5 (7.5 ; 9.4) | Reference |  |  |
| 5-6 | 323/5,366 | 6.0 (5.4 ; 6.7) | 6.8 (5.8 ; 7.8) | 0.81 | 0.70 ; 0.98 | 0.0063 |
| 7+ | 100/1,700 | 5.9 (4.8 ; 7.1) | 7.2 (5.2 ; 9.0) | 0.78 | 0.62 ; 0.98 | 0.0317 |
| Missing | 151 |  |  |  |  |  |
| Total household monthly income |  |  |  |  |  | <0.0001 |
| <1000€ | 10/201 | 5.0 (2.4 ; 9.0) | 5.4 (1.6 ; 8.4) | 0.60 | 0.32 ; 1.15 | 0.1234 |
| 1000-1499 | 18/447 | 4.0 (2.4 ; 6.3) | 4.8 (2.2 ; 7.0) | 0.48 | 0.30 ; 0.77 | 0.0026 |
| 1500-1999 | 61/1000 | 6.1 (4.7 ; 7.8) | 8.2 (5.8 ; 10.3) | 0.76 | 0.57 ; 0.99 | 0.0411 |
| 2000-2999 | 138/2,500 | 5.5 (4.7 ; 6.5) | 7.1 (5.6 ; 8.4) | 0.67 | 0.55 ; 0.82 | <0.0001 |
| 3000-3999 | 207/3,426 | 6.0 (5.3 ; 6.9) | 7.5 (6.2 ; 8.7) | 0.77 | 0.65 ; 0.92 | 0.0034 |
| >4000 | 477/6,045 | 7.9 (7.2 ; 8.6) | 9.5 (8.4 ; 10.5) | Reference |  |  |
| Missing | 1,009 |  |  |  |  |  |
| Educational level |  |  |  |  |  | <0.0001 |
| <High-school degree | 59/1,629 | 3.6 (2.8 ; 4.7) | 5.0 (3.5 ; 6.3) | Reference |  |  |
| High-school degree or undergraduate | 349/6,032 | 5.8 (5.2 ; 6.4) | 7.8 (6.9 ; 8.7) | 1.69 | 1.27 ; 2.24 | 0.0003 |
| Graduate degree or doctorate | 464/5,646 | 8.2 (7.5 ; 9.0) | 9.3 (8.3 ; 10.2) | 2.43 | 1.84 ; 3.21 | <0.0001 |
| Missing | 1,321 |  |  |  |  |  |
| Professional activity before lockdown |  |  |  |  |  | <0.0001 |
| Student | 5/81 | 6.2 (2.0 ; 13.8) | 7.2 (0.1 ; 12.6) | 0.68 | 0.27 ; 1.68 | 0.4023 |
| Working | 741/8,309 | 8.9 (8.3 ; 9.6) | 10.5 (9.5 ; 11.4) | Reference |  |  |
| Looking for a job | 30/402 | 7.5 (5.1 ; 10.5) | 7.8 (4.7 ; 10.4) | 0.83 | 0.57 ; 1.21 | 0.3305 |
| Retired | 182/5,381 | 3.4 (2.9 ; 3.9) | 4.3 (3.5 ; 5.0) | 0.35 | 0.30 ; 0.42 | <0.0001 |
| Not working due to health conditions | 7/125 | 5.6 (2.3 ; 11.2) | 3.8 (0.5 ; 6.3) | 0.58 | 0.27 ; 1.25 | 0.1621 |
| No professional activity (housewife or husband) | 16/306 | 5.2 (3.0 ; 8.4) | 7.2 (2.3 ; 11.1) | 0.53 | 0.32 ; 0.88 | 0.0144 |
| Missing | 24 |  |  |  |  |  |
| Essential job position |  |  |  |  |  |  |
| Healthcare worker Y vs N | 60/568 | 10.6 (8.2 ; 13.4) | 11.6 (8.3 ; 14.4) | 1.62 | 1.23 ; 2.13 | 0.0007 |
| Other essential job Y vs N | 122/1,425 | 8.6 (7.2 ; 10.1) | 11.9 (9.4 ; 14.0) | 1.29 | 1.06 ; 1.58 | 0.0114 |
| Professional activity during lockdown |  |  |  |  |  | <0.0001 |
| Not working | 240/6,295 | 3.8 (3.4 ; 4.3) | 4.9 (4.1 ; 5.6) | 0.39 | 0.33 ; 0.46 | <0.0001 |
| Stopped working | 127/1,457 | 8.7 (7.3 ; 10.3) | 8.3 (6.4 ; 10.0) | 0.91 | 0.74 ; 1.12 | 0.3717 |
| Working from home, remote working | 410/4,444 | 9.2 (8.4 ; 10.1) | 11.6 (10.1 ; 13.0) | Reference |  |  |
| Partially working from home | 75/759 | 9.9 (7.9 ; 12.2) | 12.6 (9.4 ; 15.1) | 1.06 | 0.82 ; 1.38 | 0.6516 |
| Working outside home | 96/1,134 | 8.5 (6.9 ; 10.2) | 11.1 (8.7 ; 13.2) | 0.89 | 0.70 ; 1.12 | 0.3062 |
| Other | 12/242 | 5.0 (2.6 ; 8.5) | 7.8 (1.0 ; 12.6) | 0.57 | 0.32 ; 1.04 | 0.0650 |
| Missing | 297 |  |  |  |  |  |
| Smoking status before lockdown |  |  |  |  |  | <0.0001 |
| Active smoker | 98/1,750 | 5.6 (4.6 ; 6.8) | 7.1 (5.4 ; 8.7) | 0.74 | 0.59 ; 0.92 | 0.0079 |
| Ex-smoker | 353/5,973 | 5.9 (5.3 ; 6.5) | 7.1 (6.1 ; 8.0) | 0.73 | 0.63 ; 0.84 | <0.0001 |
| Non smoker | 516/6,670 | 7.7 (7.1 ; 8.4) | 8.9 (7.9 ; 9.9) | Reference |  |  |
| Missing | 235 |  |  |  |  |  |
| Alcohol use before lockdown (in g/dy) |  |  |  |  |  | 0.0821 |
| <5 | 426/5,803 | 7.3 (6.7 ; 8.0) | 8.5 (7.6 ; 9.3) | Reference |  |  |
| [5,10[ | 176/2,641 | 6.7 (5.7 ; 7.7) | 8.0 (6.5 ; 9.3) | 0.94 | 0.78 ; 1.13 | 0.4971 |
| [10,20[ | 205/2,963 | 6.9 (6.0 ; 7.9) | 8.4 (6.9 ; 9.7) | 1.03 | 0.86 ; 1.23 | 0.7380 |
| [20,30[ | 63/1,359 | 4.6 (3.6 ; 5.9) | 5.8 (3.6 ; 7.7) | 0.70 | 0.53 ; 0.92 | 0.0201 |
| ≥30 | 64/1,128 | 5.7 (4.4 ; 7.2) | 6.9 (4.5 ; 8.9) | 0.87 | 0.66 ; 1.15 | 0.3276 |
| Missing | 734 |  |  |  |  |  |
| BMI (kg/m<sup>2</sup>) |  |  |  |  |  | 0.0390 |
| <18.5 | 33/499 | 6.6 (4.6 ; 9.2) | 5.6 (3.0 ; 7.7) | 0.87 | 0.61 ; 1.25 | 0.4578 |
| [18.5; 25[ | 619/8,521 | 7.3 (6.7 ; 7.8) | 8.2 (7.5 ; 9.0) | Reference |  |  |
| [25; 30[ (overweight) | 239/3,995 | 6.0 (5.3 ; 6.8) | 8.2 (7.0 ; 9.3) | 0.83 | 0.71 ; 0.97 | 0.0191 |
| ≥30 (obese) | 82/1,409 | 5.8 (4.7 ; 7.2) | 6.8 (5.0 ; 8.5) | 0.78 | 0.61 ; 0.99 | 0.0400 |
| Missing | 204 |  |  |  |  |  |
| Chronic diseases |  |  |  |  |  | <0.0001 |
| Yes | 259/4,756 | 5.5 (4.8 ; 6.1) | 6.7 (5.7 ; 7.7) | 0.72 | 0.62 ; 0.83 | <0.0001 |
| No | 715/9,767 | 7.3 (6.8 ; 7.9) | 8.7 (7.8 ; 9.4) | Reference |  |  |
| Don't know | 8/80 | 10.0 (4.4 ; 18.8) | 9.9 (0.0 ; 16.2) | 1.46 | 0.70 ; 3.06 | 0.3138 |
| Missing | 25 |  |  |  |  |  |
| Chronic diseases (Y vs N) |  |  |  |  |  |  |
| Asthma, COPD, other respir. diseases | 91/1,534 | 5.9 (4.8 ; 7.2) | 6.5 (4.9 ; 7.9) | 0.81 | 0.64 ; 1.02 | 0.0695 |
| Diabetes | 19/481 | 4.0 (2.4 ; 6.1) | 5.8 (2.5 ; 8.5) | 0.63 | 0.39 ; 1.00 | 0.0519 |
| Hypertension | 67/1,553 | 4.3 (3.4 ; 5.5) | 4.9 (3.4 ; 6.3) | 0.59 | 0.46 ; 0.77 | <0.0001 |
| Other cardiovascular diseases | 19/451 | 4.2 (2.6 ; 6.5) | 6.7 (3.3 ; 9.4) | 0.64 | 0.40 ; 1.01 | 0.0569 |
| Cancer | 53/830 | 6.4 (4.8 ; 8.3) | 7.6 (5.3 ; 9.7) | 0.83 | 0.62 ; 1.11 | 0.1963 |
| Anxiety, depression | 30/404 | 7.4 (5.1 ; 10.4) | 7.8 (3.8 ; 11.0) | 1.08 | 0.74 ; 1.58 | 0.6955 |
| Other | 106/1,826 | 5.8 (4.8 ; 7.0) | 7.3 (5.6 ; 8.8) | 0.78 | 0.63 ; 0.97 | 0.0228 |
| Missing | 25 |  |  |  |  |  |

\* with stratification on the source cohort.

**Table 4.** multivariable analysis of factors associated with a positive ELISA-S

|  | Odds-Ratio* | 95%CI | P-value |
| --- | --- | --- | --- |
| Regions |  |  |  |
| Ile-de-France | 2.43 | 2.02 ; 2.93 | <0.0001 |
| Grand-Est | 2.24 | 1.83 ; 2.75 | <0.0001 |
| Nouvelle Aquitaine | Reference |  |  |
| Age group (years) |  |  |  |
| <40 | 1.84 | 1.49 ; 2.28 | <0.0001 |
| [40-50[ | 1.92 | 1.57 ; 2.36 | <0.0001 |
| [50-60[ | Reference |  |  |
| [60-70[ | 0.77 | 0.60 ; 0.97 | 0.0299 |
| >=70 | 0.56 | 0.42 ; 0.74 | <0.0001 |
| Gender |  |  |  |
| Male | Reference |  |  |
| Female | 1.14 | 0.99 ; 1.32 | 0.0792 |
| Household size and composition- Nb children (<18yrs) |  |  |  |
| 0 | Reference |  |  |
| 1+ | 1.30 | 1.11 ; 1.53 | 0.0014 |
| Smoking status before lockdown |  |  |  |
| Active smoker | 0.71 | 0.57 ; 0.89 | 0.0033 |
| Ex-smoker | 0.96 | 0.83 ; 1.11 | 0.5607 |
| Non smoker | Reference |  |  |

235 participants – 16 with an ELISA-S positive result were excluded from the multivariable model due to missing smoking status

## Discussion

In May-June 2020 following the first wave of the COVID-19 pandemic and the subsequent lockdown in France, the seroprevalence of SARS-CoV-2-infection was 10% to 9% in the adult population in the 2 regions with the highest rate of disease and 3% in a region with a low rate. The seroprevalence of neutralizing antibody titers ≥40 or ELISA IgG against the NP protein was half that detected by ELISA IgG against the spike protein. Seroprevalence was strongly associated with reported symptoms and nearly half of the participants who tested positive experienced symptoms of COVID-19, while 1 in 5 did not recall having any symptoms. The associations between seroprevalence and age, living with at least one child or adolescent, and smoking status were consistent across all regions.

To our knowledge this is the first study evaluate the seroprevalence of SARS-CoV-2 in the general adult population in France. The rates of seroprevalence in the 3 regions were close to the cumulative proportions of infection predicted by models at the end of the lockdown period.^26^ They were also in the range reported in similar studies in Europe.^4,5,7-9^ Half of the participants with a positive ELISA-S had an episode corresponding to the definition of a COVID-19 case, and the reported symptoms corresponded to those described in similar studies.^5,7^ One in five positive participants did not experience any symptoms from the onset of the pandemic. This was lower than in Spain^5^ or England,^7^ which was around 30%, perhaps due to different methods of data collection of symptoms. Interestingly, ELISA-NP and SN seroprevalence was strongly associated with the presence and intensity of symptoms in ELISA-S positive participants. These results are similar to studies suggesting that asymptomatic or paucisymptomatic individuals have a weaker immune response to SARS-CoV-2 infection.^2,27^

A lower seroprevalence with increasing age was reported in several population-based serological studies^4,6^ and a higher rate of possible COVID-19 with decreasing age was described in an earlier study.^16^ Although men are known to be at a higher risk of severe COVID-19, hospitalization and deaths than women,^28^ we found an association between seroprevalence and female gender in NA, which was also reported in a recent Italian study.^8^ This association was only found in the region with a lower prevalence and may be related to the specific dynamics of transmission in this area. Based on the estimated 5-day median COVID-19 incubation time and the appearance of symptoms within 12 days after infection,^29^ participants who developed a possible COVID-19 before March 23 and tested positive were potentially infected before lockdown, probably in the workplace or in the community. This could explain why we did not find any specific association with social health inequalities, while, conversely, univariable analyses showed associations between seroprevalence and working adults with higher incomes and educational levels. As in other studies, univariable analysis identified the size of the household and the number of rooms, but only living with at least one child remained associated with seroprevalence on multivariable analysis, indicating that children could play an important role in household-related transmission.^30^

Finally, active smoking was associated with a lower rate of ELISA-S or SN positive results.^7,31^ Smoking status was collected before the peak of the pandemic and thus could not have been affected by preventive behaviours in smokers. Although smoking is a risk factor for severe COVID-19 in infected patients,^32^ its role in the risk of infection remains unclear because certain components of the smoke (such as nicotine) regulate ACE2 receptor expression which is involved in SARS-CoV-2 entry into cells.^33,34^ Smoking is also known to be associated with lower serum levels of IgG, IgA or IgM,^35^ but this probably does not explain why smoking was also negatively correlated with SARS-CoV-2 RT-PCR positive results in several studies.^36,37^

Our study has several limitations. First, the primary endpoint is based on a test that does not have a 100% sensitivity and specificity. Thus, certain participants were probably misclassified. We used manufacturer-defined cutoff points for positivity, although the test performance can increased by using other positive and negative cut-off values.^38^ However, prevalence correction using these reported test

performances or by the manufacturer are not applicable to our study, since the use of capillary blood on DBS and the elution procedures do not correspond to the reported experimental conditions. To overcome this limitation, we used a statistical imputation model to estimate the performance of ELISA-S, showing a sensitivity and specificity > 97.5%. The seroprevalence levels and the risk factors identified on multivariable analysis with MI were identical to those obtained with ELISA-S, which supports the robustness of our primary results.

The second potential limitation is that the selected adult population in each region may not be representative. Certain social categories were probably under- or over-represented, and although selection and participation biases were accounted for with an appropriate weighting and raking method, our findings cannot be considered to be strictly representative of the general adult population in these regions. Nevertheless, the large number of subjects from all social categories makes it possible to draw robust conclusions on the factors associated with seroprevalence. We limited the questionnaires to detailed description of symptoms present in the past 14 days, (except for acute respiratory illness) to avoid recall bias. Thus, we may have missed symptoms related to SARS-CoV-2 infection that occurred in the last week of February when SARS-CoV-2 began spreading. On the other hand, we cannot formally associate the self-reported COVID-19 symptoms with a positive serological result and once again, misclassification may have occurred.

This study has several strengths. In particular, it is based on well-characterized general population cohorts with a very high participation rate. Moreover, serological samples were collected within 1 to 3 months after the period of intense circulation of SARS-CoV-2 and all serological tests were centralized and performed blinded to participants’ characteristics or clinical history. Several serological methods were combined, including neutralization, to improve the interpretation of seroprevalence results.

In conclusion, our study shows that the level of seroprevalence remains low in the French regions most affected by the first wave of SARS-CoV-2. Longer-term clinical and serological follow-up is needed to evaluate the duration of the humoral response, the risk of infection or re-infection, and to establish the correlates of protection - a key element in preparing for evaluation of vaccines against SARS-CoV-2.

## Data Availability

The datasets used and analyzed during the current study are available from the corresponding author on reasonable request

## Acknowledgments

The authors warmly thank all the volunteers of the Constances, E3N-E4N, and NutriNet-Santé cohorts.

We thank the staff of the Constances, E3N-E4N and NutriNet-Santé cohorts that have worked with dedication and engagement to collect and manage the data used for this study and to ensure continuing communication with the cohort participants. We thank the CEPH-Biobank staff for their adaptability and the quality of their work. In the virology department, Dr Nadège Brisbarre and the technical staff for impeccable management of samples and serological assays.

We thank Mrs Dale Roche-Leclerc for her help in editing the manuscript.

## Author Statement

Dr Carrat had full access to all the data in the study and takes responsibility for the integrity of the data and the accuracy of the data analysis.

*Study idea and design:* Carrat, de Lamballerie, Rahib, Meyer, Charles, Ancel, Bajos, Severi, Touvier, Zins.

*Data acquisition:* de Lamballerie, Rahib, Blanché, Deleuze, Severi, Touvier, Zins.

*Data analysis and interpretation:* Carrat, de Lamballerie, Meyer, Bajos, Severi, Touvier, Zins.

*Drafting of the manuscript*: Carrat

*Critical revision of the manuscript for important intellectual content:* All authors.

*Statistical analysis:* Carrat, Lapidus, Artaud, Kab, Renuy, Szabo de Edelenyi *Obtained funding:* Carrat, de LamballerieSeveri, Touvier, Zins, Bajos.

*Administrative, technical, or material support:* Rahib, Blanché, Lydié, Priet, Saba Villarroel, Fourié, Nicol, Legot, Druesne-Pecollo, Essedik, Lai, Gagliolo.

*Study supervision:* Carrat, de Lamballerie, Rahib, Lydié, Charles, Ancel, Deleuze, Severi, Touvier, Zins, Bajos

## Funding

### This study

ANR (Agence Nationale de la Recherche, #ANR-20-COVI-000, #ANR-10-COHO-06), Fondation pour la Recherche Médicale (#20RR052-00), Inserm (Institut National de la Santé et de la Recherche Médicale, #C20-26).

### Cohorts funding

The CONSTANCES Cohort Study is supported by the Caisse Nationale d’Assurance Maladie (CNAM), the French Ministry of Health, the Ministry of Research, the Institut national de la santé et de la recherche médicale. CONSTANCES benefits from a grant from the French National Research Agency [grant number ANR-11-INBS-0002] and is also partly funded by MSD, AstraZeneca, Lundbeck and L’Oreal.

The E3N-E4N cohort is supported by the following institutions: Ministère de l’Enseignement Supérieur, de la Recherche et de l’Innovation, INSERM, University Paris-Saclay, Gustave Roussy, the MGEN, and the French League Against Cancer. The NutriNet-Santé study is supported by the following public institutions: Ministère de la Santé, Santé Publique France, Institut National de la Santé et de la Recherche Médicale (INSERM), Institut National de la Recherche Agronomique (INRAE), Conservatoire National des Arts et Métiers (CNAM) and Sorbonne Paris Nord. The CEPH-Biobank is supported by the « Ministère de l’Enseignement Supérieur, de la Recherche et de l’Innovation ».

## The SAPRIS study group

Nathalie Bajos (co-Principal investigator), Fabrice Carrat (co-Principal investigator), Pierre-Yves Ancel, Marie-Aline Charles, Florence Jusot, Claude Martin, Laurence Meyer, Ariane Pailhé, Gianluca Severi, Alexis Spire, Mathilde Touvier, Marie Zins.

## The SAPRIS-SERO study group

Fabrice Carrat (Principal investigator), Pierre-Yves Ancel, Marie-Aline Charles, Gianluca Severi, Mathilde Touvier, Marie Zins

Sofiane Kab, Adeline Renuy, Stephane Le-Got, Celine Ribet, Emmanuel Wiernik, Marcel Goldberg, Marie Zins (Constances cohort),

Fanny Artaud, Pascale Gerbouin-Rérolle, Mélody Enguix, Camille Laplanche, Roselyn Gomes-Rima, Lyan Hoang, Emmanuelle Correia, Alpha Amadou Barry, Nadège Senina, Gianluca Severi (E3N-E4N cohort)

Fabien Szabo de Edelenyi, Nathalie Druesne-Pecollo, Younes Esseddik, Serge Hercberg, Mathilde Touvier (NutriNet-Santé cohort)

Marie-Aline Charles, Pierre-Yves Ancel, Valérie Benhammou, Anass Ritmi, Laetitia Marchand, Cecile Zaros, Elodie Lordmi, Adriana Candea, Sophie de Visme, Thierry Simeon, Xavier Thierry, Bertrand Geay, Marie-Noelle Dufourg, Karen

Milcent (Epipage2 and Elfe child cohorts)

Clovis Lusivika-Nzinga, Gregory Pannetier, Nathanael Lapidus, Isabelle Goderel, Céline Dorival, Jérôme Nicol, Fabrice Carrat (IPLESP – methodology and coordinating data center)

Cindy Lai, Hélène Esperou, Sandrine Couffin-Cadiergues (Inserm)

Jean-Marie Gagliolo (Institut de Santé Publique)

Hélène Blanché, Jean-Marc Sébaoun, Jean-Christophe Beaudoin, Laetitia Gressin, Valérie Morel, Ouissam Ouili, Jean-François Deleuze (CEPH-Biobank)

Stéphane Priet, Paola Mariela Saba Villarroel, Toscane Fourié, Souand Mohamed Ali, Abdenour Amroun, Morgan Seston, Nazli Ayhan, Boris Pastorino, Xavier de Lamballerie (Unité des Virus Emergents)

## References

1. Koopmans M, Haagmans B. Assessing the extent of SARS-CoV-2 circulation through serological studies. Nat Med 2020; 26(8): 1171–2.

2. Azziz NH, Corman VM, Echterhoff AKC, et al. Seroprevalence and correlates of SARS-CoV-2 neutralizing antibodies: results from a population-based study in Bonn, Germany. MedRxiv 2020; preprint. doi:2020.08.24.20181206.

3. Gudbjartsson DF, Norddahl GL, Melsted P, et al. Humoral Immune Response to SARS-CoV-2 in Iceland. N Engl J Med 2020. Sep 1. Online ahead of print.

4. Stringhini S, Wisniak A, Piumatti G, et al. Seroprevalence of anti-SARS-CoV-2 IgG antibodies in Geneva, Switzerland (SEROCoV-POP): a population-based study. Lancet 2020. 396(10247):313–9.

5. Pollan M, Perez-Gomez B, Pastor-Barriuso R, et al. Prevalence of SARS-CoV-2 in Spain (ENE-COVID): a nationwide, population-based seroepidemiological study. Lancet 2020. 396(10250):535–44.

6. UK Biobank SARS-CoV-2 Serology Study. Weekly Report - 21 July 2020. https://www.ukbiobank.ac.uk/2020/07/uk-biobank-covid-19-antibody-study-latest-updates/ (accessed August 28th, 2020).

7. Ward H, Atchinson C, Whitaker M, et al. Antibody prevalence for SARS-CoV-2 following the peak of the pandemic in England: REACT2 study in 100,000 adults. MedRxiv 2020. preprint. doi:2020.08.12.20173690.

8. Vena A, Berruti M, Adessi A, et al. Prevalence of Antibodies to SARS-CoV-2 in Italian Adults and Associated Risk Factors. J Clin Med 2020; 9(9): E2780.

9. Herzog S, De Bie J, Abrams S, et al. Seroprevalence of IgG antibodies against SARS coronavirus 2 in Belgium: a prospective cross-sectional nationwide study of residual samples. MedRxiv 2020. preprint. doi:2020.06.08.20125179.

10. Xu X, Sun J, Nie S, et al. Seroprevalence of immunoglobulin M and G antibodies against SARS-CoV-2 in China. Nat Med 2020; 26(8): 1193–5.

11. Silveira MF, Barros AJD, Horta BL, et al. Population-based surveys of antibodies against SARS-CoV-2 in Southern Brazil. Nat Med 2020; 26(8): 1196– 9.

12. Skowronski DM, Sekirov I, Sabaiduc S, et al. Low SARS-CoV-2 sero-prevalence based on anonymized residual sero-survey before and after first wave measures in British Columbia, Canada, March-May 2020. MedRxiv 2020. preprint. doi:2020.07.13.20153148.

13. Havers FP, Reed C, Lim T, et al. Seroprevalence of Antibodies to SARS-CoV-2 in 10 Sites in the United States, March 23-May 12, 2020. JAMA Intern Med 2020. Jul 21. Online ahead of print.

14. Sood N, Simon P, Ebner P, et al. Seroprevalence of SARS-CoV-2-Specific Antibodies Among Adults in Los Angeles County, California, on April 10-11, 2020. JAMA 2020. 323(23):2425–7.

15. Menachemi N, Yiannoutsos CT, Dixon BE, et al. Population Point Prevalence of SARS-CoV-2 Infection Based on a Statewide Random Sample - Indiana, April 25-29, 2020. MMWR Morb Mortal Wkly Rep 2020; 69(29): 960–4.

16. Carrat F, Touvier M, Severi G, et al. Incidence and risk factors of illnesses presumably caused by a SARS-CoV-2 infection in the general population during the lockdown period: a multi-cohort study. submitted 2020;

17. Clavel-Chapelon F, Group ENS. Cohort Profile: The French E3N Cohort Study. Int J Epidemiol 2015; 44(3): 801–9.

18. Hercberg S, Castetbon K, Czernichow S, et al. The Nutrinet-Sante Study: a web-based prospective study on the relationship between nutrition and health and determinants of dietary patterns and nutritional status. BMC Public Health 2010; 10: 242.

19. Zins M, Goldberg M, team C. The French CONSTANCES population-based cohort: design, inclusion and follow-up. Eur J Epidemiol 2015; 30(12): 1317–28.

20. Gallian P, Pastorino B, Morel P, Chiaroni J, Ninove L, de Lamballerie X. Lower prevalence of antibodies neutralizing SARS-CoV-2 in group O French blood donors. Antiviral Res 2020; 181:104880.

21. Santé Publique France. COVID-19 : point épidémiologique du 14 mai 2020, 2020. https://www.santepubliquefrance.fr/content/download/252588/2603686 (accessed August 24th 2020).

22. Obesity: preventing and managing the global epidemic. Report on a WHO consultation. WHO. Geneva, 1997.

23. European Center for Diseases Control. Case definition for coronavirus disease 2019 (COVID-19), as of 29 May 2020. https://www.ecdc.europa.eu/en/covid-19/surveillance/case-definition (accessed June 15th 2020).

24. Deville JC, Sarndal CE, Sautory O. Generalized Raking Procedures in Survey Sampling. J Am Stat Assoc 1993; 88(423): 1013–20.

25. Rubin DB. Multiple Imputation for Nonresponse in Surveys. New York: John Wiley & Sons; 1987.

26. Salje H, Tran Kiem C, Lefrancq N, et al. Estimating the burden of SARS-CoV-2 in France. Science 2020; 369(6500): 208–11.

27. Long QX, Tang XJ, Shi QL, et al. Clinical and immunological assessment of asymptomatic SARS-CoV-2 infections. Nat Med 2020; 26(8): 1200–4.

28. Williamson EJ, Walker AJ, Bhaskaran K, et al. Factors associated with COVID-19-related death using OpenSAFELY. Nature 2020; 584(7821): 430–6.

29. Lauer SA, Grantz KH, Bi Q, et al. The Incubation Period of Coronavirus Disease 2019 (COVID-19) From Publicly Reported Confirmed Cases: Estimation and Application. Ann Intern Med 2020; 172(9): 577–82.

30. Jing QL, Liu MJ, Zhang ZB, et al. Household secondary attack rate of COVID-19 and associated determinants in Guangzhou, China: a retrospective cohort study. Lancet Infect Dis 2020. Jun 17. Online ahead of print.

31. Fontanet A, Tondeur L, Madec Y, et al. Cluster of COVID-19 in northern France: A retrospective closed cohort study. MedRxiv 2020. preprint. doi:2020.04.18.20071134.

32. Guan WJ, Ni ZY, Hu Y, et al. Clinical Characteristics of Coronavirus Disease 2019 in China. N Engl J Med 2020; 382(18): 1708–20.

33. Oakes JM, Fuchs RM, Gardner JD, Lazartigues E, Yue X. Nicotine and the renin-angiotensin system. Am J Physiol Regul Integr Comp Physiol 2018; 315(5): R895–R906.

34. Smith JC, Sausville EL, Girish V, et al. Cigarette Smoke Exposure and Inflammatory Signaling Increase the Expression of the SARS-CoV-2 Receptor ACE2 in the Respiratory Tract. Dev Cell 2020; 53(5): 514–29 e3.

35. Gulsvik A, Fagerhoi MK. Smoking and immunoglobulin levels. Lancet 1979; 1(8113): 449.

36. Rentsch CT, Kidwai-Khan F, Tate JP, et al. Covid-19 Testing, Hospital Admission, and Intensive Care Among 2,026,227 United States Veterans Aged 54-75 Years. medRxiv 2020. preprint. doi:2020.04.09.20059964.

37. Simons D, Shahab L, Brown J, Perski O. The association of smoking status with SARS-CoV-2 infection, hospitalisation and mortality from COVID-19: A living rapid evidence review with Bayesian meta-analyses (version 7). Qeios 2020. https://www.qeios.com/read/UJR2AW.8 (accessed Sep 10th, 2020).

38. Meyer B, Torriani G, Yerly S, et al. Validation of a commercially available SARS-CoV-2 serological immunoassay. Clin Microbiol Infect 2020. Jun 27. nline ahead of print.

